# Quasi-experimental evaluation of a social media campaign to promote HPV vaccination in Bauchi state, Nigeria

**DOI:** 10.64898/2026.11.11.26367870

**Authors:** William Douglas Evans, Alec Ulasevich, Ibrahim Sani Mohammed Gimba, Sarah Francis, Julie Szabo, Sean O’Sullivan, Drew Bernard

## Abstract

Cervical cancer is the leading cause of cancer death in women in 26 countries, including in sub-Saharan African countries. Cervical and other cancer risks can be greatly reduced through timely HPV vaccination and early screening. But it continues to pose a significant public health challenge in LMICs such as Nigeria. This study reports on the implementation and evaluation of a pair of social media behavior change campaigns to increase HPV vaccination amongst girls ages 9-14 and reduce vaccine hesitancy amongst girls and their parents in Bauchi state, Nigeria. The primary objectives are to conduct a quasi-experimental evaluation of the social media campaign; to examine the effect of higher versus lower levels of campaign exposure on HPV vaccination and related outcomes; to determine whether the social media campaign followed its planned theory of change; and to identify which social media messages were most effective in promoting HPV vaccination.

We implemented an HPV vaccination social media promotion campaign, using videos designed to encourage parents to vaccinate their daughters, at scale in Bauchi state on the Meta and TikTok social media platforms. We used a quasi-experimental design, a heavy-up approach, that experimentally manipulated the level of social media content delivered in Bauchi. We used the Virtual Lab platform, an open-source platform that uses social media to recruit participants using advertising and creates panels of respondents for data collection through an online survey, and also collected social media metrics including impressions and engagement. The campaign was effective in increasing HPV vaccination rates from 69% at baseline to 86% at endline over 2 campaign phases that spanned approximately 3 months. Based on GEE modeling, we found that higher levels of engagement with the campaign were associated with higher vaccination rates at midline and endline. Pro-vaccine attitudes and reduced vaccine hesitancy also predicted higher vaccination rates. Also, the campaign produced additional HPV vaccinations at low cost: Adjusted for campaign exposure cost of $0.81 per additional vaccination in campaign phase 1, $2.10 per vax in phase 2, and $2.40 for the total campaign period.

Evidence from this study indicates that social media may be an effective strategy to promote HPV vaccination at low cost. Future rigorous trials using experimental methods are needed to further evaluate social media to promote HPV vaccination.

## INTRODUCTION

The World Health Organization (WHO) reports that cervical cancer is the fifth most commonly diagnosed cancer in women, with 604,000 new cases estimated in 2024, and that it is the leading cause of cancer death in women in 26 countries, including in sub-Saharan Africa.^1^ Lack of systemic Human Papillomavirus (HPV) vaccination and screening are a significant contributor to the burden of cancer among women in low and middle income countries (LMIC) generally, and sub-Saharan Africa in particular,^2^ unlike in many developed countries. A report by the WHO ranks Nigeria fifth globally in cervical cancer deaths, with estimates of 12,000 new cases and 8,000 deaths annually.^3^

Cervical and other cancer risks can be greatly reduced through timely HPV vaccination and early screening. But it continues to pose a significant public health challenge in LMICs such as Nigeria.^4^ Risk factors include lack of knowledge about the benefits of HPV vaccination, concerns about safety and efficacy of the vaccine, and mis- and disinformation about health risks.^5^ Interventions to promote acceptance among parents and adolescent girls and young women, and to increase motivation, opportunity, and ability to receive the HPV vaccine are needed.^6,7^

Social media has been studied as a potentially promising approach to promote HPV vaccination and reduce vaccine hesitancy.^8^ Social media content can potentially promote the benefits of vaccination, attempt to reduce barriers (such as myths and misinformation about health risks), and increase acceptance and intentions to vaccinate.^9^ Social media content can use strategies such as influential community members,^10^ entertainment education,^11^ and raise awareness about opportunities and ability to vaccinate.^12^ However, there have been relatively few studies that have rigorously evaluated the efficacy of social media to promote HPV vaccination.^13^ More research using experimental and quasi-experimental methods are needed in this area.

Nigeria has increased efforts in recent years to increase multiple forms of community health promotion and healthcare, including vaccination, in recent years.^14^ These include promotion efforts such as through Maternal, Newborn, and Child Health Week (MNCH) efforts led by the Ministry of Health (MoH).^15^ One potential opportunity to increase HPV vaccination is to promote MNCH week participation, and to encourage parents to bring their adolescent girls to receive the HPV vaccine at these events. Social media may be an effective strategy to achieve this goal.

### Social Media Campaign to Promote HPV Vaccination in Bauchi State Nigeria

This study reports on the implementation and evaluation of a pair of social media behavior change campaigns to increase HPV vaccination amongst girls ages 9-14 and reduce vaccine hesitancy amongst girls and their parents in Bauchi state, Nigeria. Bauchi is the fifth largest state in the country, located in the northern and central part of Nigeria, with a population of over 8.3 million in 2025.^16^ This evaluation study, funded by the Gates Foundation, has the overall objective of identifying which campaigns and strategies are effective in increasing HPV vaccination rates and reducing vaccine hesitancy, and at what level of cost-effectiveness.

The current study analyzed a theory of change (ToC) co-developed with the campaign implementation team in order to understand the processes and effects of social media in influencing HPV vaccination and vaccine hesitancy.^17^ The ToC posited that social media could influence drivers of vaccine hesitancy, according to the 5 Cs approach, by promoting motivation, opportunity, and ability to vaccinate, and by promoting pro-vaccination social norms.^18,19,20^

The Gates Foundation’s primary goal for the HPV vaccine is to achieve the WHO target of eliminating cervical cancer by ensuring 90% of girls worldwide are vaccinated against HPV by age 15.^21^ The foundation supports this by funding initiatives with multi-lateral organizations and major non-governmental organizations (NGOs) to increase vaccine access, support research into single-dose vaccines and new HPV vaccines, and partner with governments and organizations to scale up vaccination programs in low- and middle-income countries. There is a specific focus on increasing HPV vaccination rates, and reducing vaccine hesitancy, among 9-14 year-old girls in priority states in Nigeria, including Bauchi state. As part of this initiative, our research team sought to evaluate the efficacy of social media to increase HPV vaccination rates and reduce vaccine hesitancy in Bauchi state.

### Specific objectives of the study

The primary objectives to be evaluated in this study are: 1) To conduct a quasi-experimental evaluation of a social media campaign to encourage parents to attend the MNCH week community events in Bauchi state and have their 9-14 year old girls vaccinated for HPV; 2) To examine the effect of higher versus lower levels of campaign exposure on HPV vaccination and related outcomes including social norms and vaccine hesitancy; 3) Determine whether the social media campaign followed its planned theory of change; 4) Identify which social media messages were most effective in promoting vaccination at the MNCH week events and in reducing vaccine hesitancy.

## METHODS

### Study Design

This study employed a quasi-experimental design in which we compared the effects of higher versus lower levels of paid advertising (boosted) social media campaign activity on HPV vaccination outcomes. This approach, known as a “heavy-up” design in advertising research, has been widely used, including studies in health promotion and public health.^22,23^

### Ethical consideration

This research, including procedures, instruments, and informed consent form, was reviewed and approved by the National Health Research Ethics Committee (NHREC) of Nigeria and the Bauchi State Ministry of Health on 7 May 2026.

Participants were contacted through the Meta and TikTok social media platforms through a paid advertising campaign. Interested participants clicked on a link and were taken to an informed consent form that provided details on the study procedures, delivered through the Facebook Messenger app. By clicking on an “accept” link after reading the consent, they were entered into a screening protocol for the study. Participants who were parents of girls age 9-14 and lived in Bauchi state were eligible and offered the survey.

Data collection began on 22 May 2026. Over the following 4 weeks prior to MNCH week, in May and June 2026, we collected survey data from a baseline sample of parents of girls ages 9-14 in Bauchi. Then we ran the initial social media campaign (phase 1) at a defined level of boosted campaign activity for approximately 4 weeks. Following this campaign, MNCH week promoting HPV vaccination occurred in early July 2026. We then collected a 2^nd^ round of survey data (midline) from the same parents. Then we ran the campaign at approximately 3 times the level of boosted activity (3 times the advertising spending to generate exposure to content), which included the MNCH week events. After the end of MNCH week, we collected midline data from the same sample of parents. We then ran the phase 2 campaign to promote vaccination at other events and activities in Bauchi state in July and August 2026. The endline survey was conducted immediately after the end of phase 2 in August 2026.

### Intervention

The Bauchi Ministry of Health and local partners, in collaboration with our investigative team, developed social media content in English and Hausa. Content focused on the benefits of HPV vaccination, and used, in different social media videos, techniques including story telling (narratives), humor, and music.

Attendance at MNCH and opportunities to vaccinate were promoted. Trusted voices from across Bauchi State were featured in the social media. Health officials, imams, doctors, mothers, local journalists, poets and comedians all carried the same message: Bring your daughter for a free HPV vaccine. In-person outreach events like MNCH Week then turned that intent into action, with free vaccines, free medical care and giveaways at health facilities across the state.

We used a social media campaign management and data collection platform called *Virtual Lab* to support study implementation. Virtual Lab was managed by the study team and used to recruit a panel of adult parent participants living in the Bauchi. The panel was recruited using Meta platform (Facebook and Instagram) paid posts using targeting persona-specific campaigns to survey respondents via remarketing and measure impacts. We collected a baseline sample of 2,206 individual Bauchi parents who met inclusion criteria.

### Measures

An online, self-administered survey delivered via the Virtual Lab platform was used for data collection in each identified state. The survey was designed for completion using Facebook Messenger software (chat feature in which each individual chat contains a survey question), and required approximately 15 minutes to complete at each time point. We offered participants incentives of 1500 Naira in mobile phone credit per completed survey to maximize completion rates.

Evaluation measures included: demographics (e.g., age, gender, education, work role, income); psychographics (e.g., psychological traits, values, lifestyle variables); vaccination social norms; related vaccine/vaccination beliefs; vaccination history; acceptance/uptake of HPV vaccination; use of social media; exposure to vaccine-specific information (on social media and other sources). Measures were derived from previous studies by the investigators, and published in the peer-reviewed literature.^18,20^

### Cost-effectiveness evaluation

Behavior change interventions that rely on social networks for their success are hypothesized to have greater impact than interventions that rely on changes in individual behavior.^24^ Thus, in addition to the effect of treatment-on-the-treated, social network researchers have measured the effect of treatment on the untreated. Researchers who have evaluated door-to-door campaigns to get out the vote, for example, have measured a smaller, discernable effect of such campaigns on the untreated (i.e., effects on members of the household who are not directly exposed to the canvasser). This additive effect may be due to effects on social norms and other social influences (e.g., based on observation of those who receive treatment by those who do not). This has implications for cost-effectiveness of social network interventions. This evaluation examined these potential effects.

The cost to implement a social media campaign to reduce vaccine hesitancy is a key input for future decision-makers considering broader implementation of the strategy following this effectiveness trial. The goal of the cost-effectiveness analysis component of the evaluation was to quantify the societal costs of implementing the social media campaign and to evaluate the cost-effectiveness of the intervention compared to the current standard of care.

We used a micro-costing approach to evaluate resources utilized for all program-related activities using a standard three-stage process of resource identification, measurement, and valuation.^25^ Costs are presented in US Dollars using the most recent Purchasing Power Parity (PPP) conversion factors at the time of the analysis.

The development of a model-based evaluation is beyond the scope of this project; we used a time horizon for the analysis consistent with the intervention time period. The intention-to-treat population was used for the primary cost-effectiveness analysis of the incremental cost-effectiveness ratio (ICER) for the cost per HCP vaccinated at follow-up. We used bootstrap sampling with replacement from intervention and control groups to estimate 95% uncertainty intervals around ICERs.^26^

### Sample size determination

We conducted an individual-level power analysis using HPV vaccination rate as the outcome of interest. Based on this analysis, we concluded that the total sample size required to achieve statistical power of 0.8 for testing the null hypothesis of no difference between the high exposure and low exposure study conditions in self-reported HPV vaccination at a given point in time (follow-up) using a two-sided test resulted in a required sample size of 2,000. Our final collected baseline sample was 2,206, exceeding power requirements.

### Sampling strategy

The sample was drawn using the Virtual Lab procedures described below. We sampled from Facebook users who meet eligibility criteria of being a parent with a girl age 9-14 living at home in Bauchi state.

### Data collection procedure

Data collection was conducted by Virtual Lab. The Virtual Lab is an open-source platform that uses social media such as Facebook and Instagram to recruit participants using advertising and creates panels of respondents for data collection. The data collection procedures may be summarized as follows.

All data in flight is encrypted via TLS and all data at rest is encrypted on encrypted block storage provided by Google Cloud Services. Data downloaded by researchers from the web portal is encrypted via TLS to their local machines at which point they must store all data which might contain personal information on encrypted disks. Respondents can request to see or delete their data at any time by sending an email to an address which is provided to them. Data deletion requests are handled within 30 days and fully removed from data archives in 60 days after the handling of the deletion request.

Participants were recruited via an ad that is shown both on Facebook and Instagram. The ad advertised that we want them to take a survey and also explains that they will receive a mobile credit as compensation for their participation.

Ads were targeted to potential respondents via Meta (Facebook and Instagram) targeting demographics including the geographic region in which they live, the industry in which they work (eg, healthcare), their job title, their education, their interests, their age, and their gender. This targeting was used in the study to ensure representativeness across the desired variables.

On clicking the ad, respondents were directed to a chat window within Facebook Messenger (FM). This chat window had an initial message which explains again that we would like them to participate in a study and that they will receive mobile credit for participating. They had the option to click "start" in the chat to start the survey.

Once the respondent clicked "start", Meta sent the Virtual Lab platform a notification that a user with a page-scoped user ID (account) number had started a chat. This ID was used to identify the user within the Virtual Lab platform and used to communicate with Facebook. The ID was usable for tracking all user interaction within the chat window and with the page more broadly, but not usable to track user interaction with any other page (thus, it is "page-scoped").

The page-scoped user id was used for "remarketing", by allowing "custom audiences" of 100+ ID’s to be created and used for ad targeting. We used remarketing to test the impact of ad campaigns on "treated" groups of individuals.

For follow-up data collection, participants were sent a message through FM indicating that a new survey was available. Repeat messages were sent for 7 days until the respondent either completed the survey, or the participant was deemed lost to follow-up.

### Data Analysis

We used both the survey data and quantitative social media metrics (reach, engagement) data drawn from the Meta platform, capturing views and interactions (likes, comments, shares) with the campaign HPV vaccination posts. Social media metrics characterize prevention message dissemination, reach, and network engagement, and provide an exogenous measure of participant exposure to content. Our analysis aimed to isolate the effects of exposure on vaccination and related attitudes and beliefs. SPSS version 31 was used for all analyses.^27^

Based on baseline data, we conducted reliability analyses for all potential scales. In addition, also using baseline data we used confirmatory factor analysis (CFA) to assess the dimensionality and construct validity of the various scales. The CFA used principal component analysis, varimax rotation and eigen value of one was the threshold to retain a factor. CFA yielded a four-factor solutions. The factors resulting from this analysis are summarized in Table 4.

- Attitudes toward HPV: feelings about and perceived importance of vaccinating one’s daughter
- Social norms: Estimates of the percentages of people close to one, and the overall population of Nigeria who vaccinate their daughters as well percentages of health care professionals who recommend vaccination
- Social influence: perceived beliefs of family and friends regarding importance of HPV vaccination
- Vaccine hesitancy factor: Set of questions representing perceived barriers to and negative believes about HPV vaccines

A summary score for each factor was calculated by taking the average of the items comprising each factor and had factor loading of at least .05. When necessary, individual items were reverse coded so that higher value represented a pro vaccine sentiment.

The study used a longitudinal design. Thus for the analyses of the campaign effects, only the participants who completed all three waves of data collection (Baseline, Midline and Endline) were included. The final total number participants was 546. Univariate repeated measures analyses were conducted as manipulation checks for the effects of campaign phase on attitude measures, self-reported exposure and vaccine uptake.

The campaign theory of change posits that engagement in the campaign predicts intervention effects on vaccination intentions and uptake. We calculated a social media engagement variable for use in Generalized estimating equations (GEE) models as an independent variable to capture the effects of the heavy-up design increase in campaign exposure between campaigns 1 and 2. Attitude measures as defined factor summary scores as well as self-reported exposure were key covariates.

## RESULTS

SPSS version 30 was used for all statistical analyses. Table 1 provides a description of the final analytic sample. Overall, about 66% of respondents self-reported as male, over 77% were under 40 years of age, over 81% reported being Muslim, and over 53% reported having a Bachelor’s Degree of higher education.

**Table 1.** Demographic Characteristics of the analysis sample.

| **Variables** | **N** | **Percent (%)** |
|---|---|---|
| **Gender** |  |  |
| Woman | 179 | 32.8% |
| Man | 360 | 65.9% |
| Non-binary or transgender | 2 | 0.4% |
| Prefer not to say | 5 | 0.9% |
| **Age** |  |  |
| 18-29 years | 196 | 35.9% |
| 30-39 years | 227 | 41.6% |
| 40-49 years | 97 | 17.8% |
| 50-54 years | 26 | 4.8% |
| **Religion** |  |  |
| Catholic | 23 | 4.2% |
| Other Christian | 76 | 13.9% |
| Islam | 444 | 81.3% |
| Other | 3 | 0.5% |
| Education |  |  |
| Primary school | 2 | 0.4% |
| Secondary school | 72 | 13.2% |
| Diploma | 161 | 29.5% |
| Bachelors | 244 | 44.7% |
| Masters | 41 | 7.5% |
| PhD | 7 | 1.3% |
| Other | 19 | 3.5% |
| **Employment** |  |  |
| Full time | 156 | 28.6% |
| Part time | 96 | 17.6% |
| Own a business | 211 | 38.7% |
| Not employed | 82 | 15.0% |

Table 2 summarizes Meta platform social media metrics for campaigns 1 and 2. Overall, the three-second and full views of campaign content were significantly higher at endline, consistent with the heavy-up design which sought to substantially increase exposure during campaign 2 compared to campaign 1.

**Table 2.**
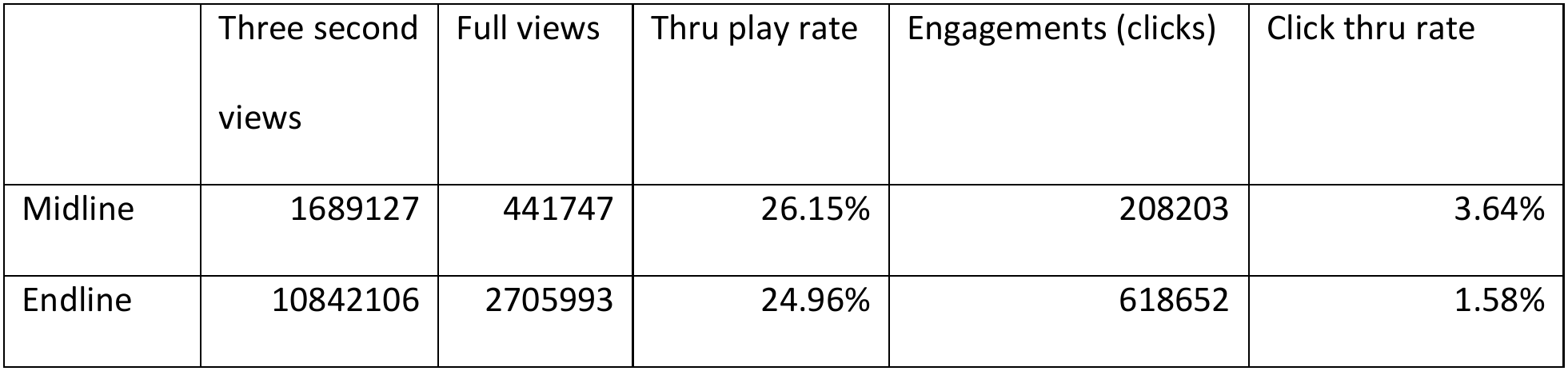
Social media metrics.

|  | Three second views | Full views | Thru play rate | Engagements (clicks) | Click thru rate |
|---|---|---|---|---|---|
| Midline | 1689127 | 441747 | 26.15% | 208203 | 3.64% |
| Endline | 10842106 | 2705993 | 24.96% | 618652 | 1.58% |

Table 3 summarizes self-reported exposure to HPV vaccination content on social media at baseline, and after campaign 1 and campaign 2. Overall, significantly more participants reported seeing HPV information at midline compared to baseline (p=0.008). Also, significantly more participants reported seeing HPV information at endline than midline (p=.007). The campaigns reached participants and they demonstrated recall of the HPV vaccination promotion content.

**Table 3.** Self-reported social media exposure.

|  | Baseline | Midline | Endline |
|---|---|---|---|
| Yes | 69.05% | 75.09% | 80.59% |
| No, I did not use this channel | 6.96% | 4.76% | 5.31% |
| No, used this channel but did not see information on HPV | 16.85% | 12.82% | 8.79% |
| Do not remember | 7.14% | 5.13% | 3.48% |

Table 4 shows results for the vaccine-related attitudes and beliefs based on summary scores obtained from factor analysis. Analyses of the significant main effects (Pro-vaccine Attitudes and Social Norms) showed that Endline scores were significantly higher than both Baseline and Midline scores (p.>.001). There was no difference between the baseline and midline scores. Analysis of the trend for Social Influence scores showed the same pattern: Endline scores were significantly higher than both baseline and midline scores.

**Table 4.** Summary factor scores and vaccine uptake by wave.

|  | Pro-Vaccine Attitude | Social Norms | Social Influence | Vaccine Hesitancy | Vaccine uptake |
|---|---|---|---|---|---|
| Baseline | 4.66 | 3.62 | 3.90 | 3.72 | 69% |
| Midline | 4.67 | 3.65 | 3.86 | 3.74 | 83% |
| Endline | 4.72 | 3.78 | 3.96 | 3.77 | 86% |

Table 4 also shows HPV vaccination rates by study time point. These rates are in response to the survey question: “Has your daughter received an HPV vaccine?” Overall, preliminary analyses using the McNemar test^28^ showed significant increases in the vaccination rates: Higher vaccination rate at midline in comparison to Baseline (p<.001); higher vaccination rate at endline in comparison to midline (p<.001).

Table 5 presents the GEE models that compared HPV vaccination rates between baseline and endline. The independent variable in these models was the campaign exposure variable based on reported social media engagement based on Meta platform campaign reach metrics. Baseline was coded with 0 since the campaign was not implemented at that time. Midline was coded as 1. Endline value was a ratio of endline engagements over midline engagement, a ratio signifying an increase in the intensity of the campaign (3.1 higher in phase 2). Individual level variables: self-reported exposure to campaign ads and attitude measures were entered as a covariate. Given the dichotomous outcome variable, we used a used logit model with binary distribution.

**Table 5.** GEE Models Predicting HPV Vaccination Rates.

| Parameter | B | Std. Error | 95% Wald Confidence Interval |  | Hypothesis Test |  |  |
|---|---|---|---|---|---|---|---|
|  |  |  | Lower | Upper | Wald Chi-Square | df | Sig. |
| (Intercept) | 0.77 | 0.0925 | 0.589 | 0.952 | 69.309 | 1 | <.001 |
| **Social media engagement** | **0.113** | **0.0299** | **0.055** | **0.172** | **14.299** | **1** | **<.001** |
| Self-reported of exposure to campaign ads | 0.016 | 0.0173 | -0.018 | 0.05 | 0.869 | 1 | 0.351 |
| **Pro vaccine attitude (Factor 1)** | **0.195** | **0.0572** | **0.083** | **0.307** | **11.613** | **1** | **<.001** |
| Vaccine hesitance (Factor 2) | -0.06 | 0.057 | -0.172 | 0.052 | 1.108 | 1 | 0.293 |

Overall, the engagement variable was a significant predictor of vaccination, related to higher vaccination rates at Midline and Endline (p<.001). Higher engagement predicted higher rates of vaccination. Among individual level variables pro-vaccine attitude (see Table 4) had significant positive effect on vaccination rates: More positive pro-vaccine attitudes were associated with higher vaccination rates at Midline and Endline (p<.001).

Tables 6 and 7 present findings from the cost effectiveness analysis. Table 7 summarizes the cost per person for the two campaigns (phases 1 and 2) and the total campaign effort. Reach represents the number of unique social media users that viewed the campaign. The cost per person was the corresponding cost per phase divided by total reach. Overall, the cost per person was $0.36 to $0.38 per person in the two phases, and the overall cost was $1.37 for the total effort. The phase 2 cost was slightly higher in phase 2 due to the heavy-up approach, and the total campaign cost was $409,821.41.

**Table 6.** Cost Effectiveness Summary.

|  |  |
|---|---|
| Reach Phase One (Midline) | 546,786 |
| Reach Phase Two (Endline) | 560,768 |
| Cost Phase One | \$197,434.70 |
| Cost Phase Two | \$212,386.72 |
| Cost Campaign | \$409,821.41 |
| Cost per person Phase One | \$0.36 |
| Cost per person Phase Two | \$0.38 |
| Cost per person Campaign | \$1.37 |

**Table 7.**
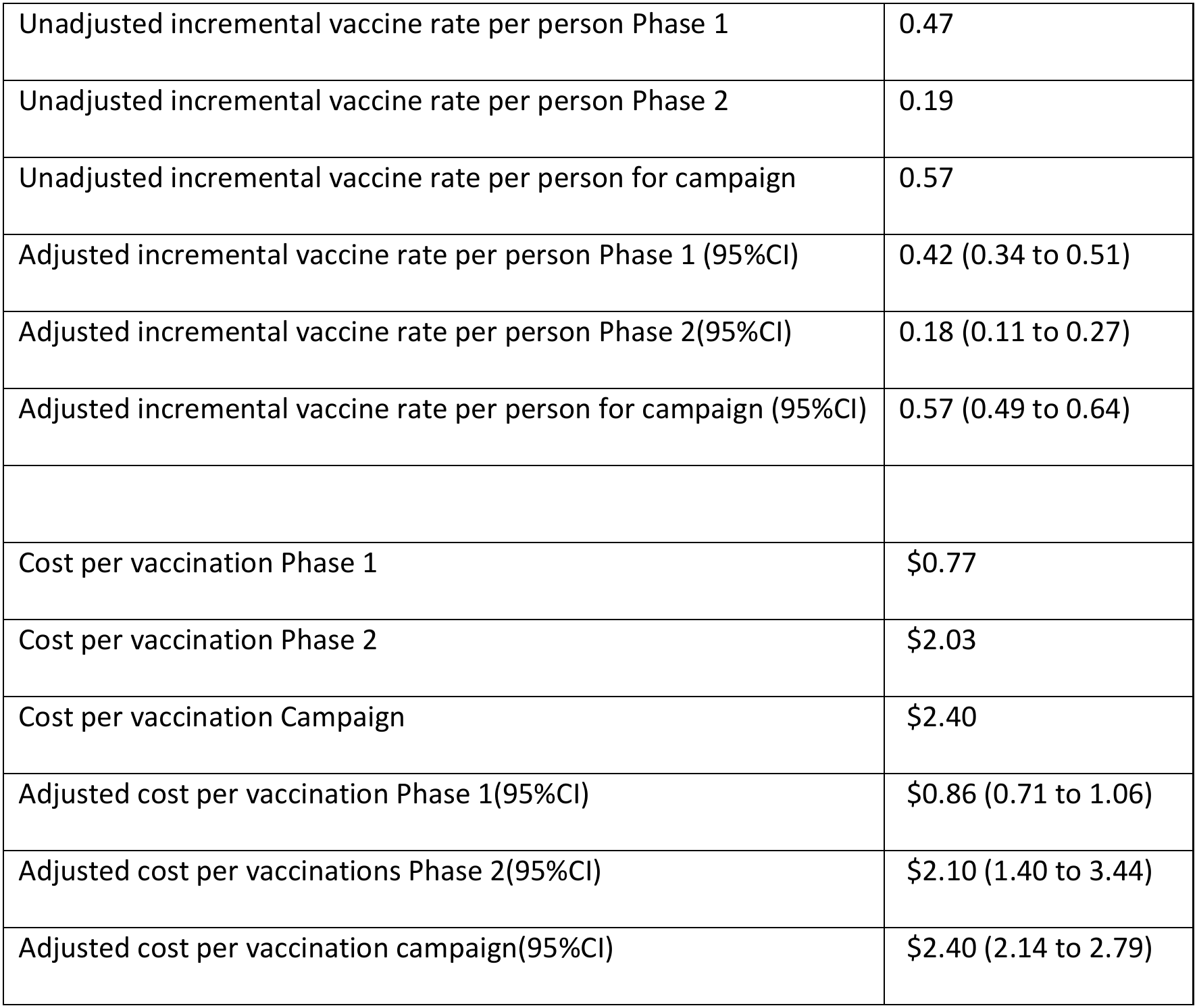
Cost of the Campaigns per Vaccination.

Table 8 summarizes the cost of the campaigns per additional vaccination. The cost per vaccination is the ratio of the cost per person over incremental vaccine rate per person. The estimated cost per vaccine was lower in phase 1 than in phase 2. The overall cost for a new vaccination for the campaign falls within the confidence interval for the adjusted cost per new vaccine for phase 2.

## DISCUSSION

In this project, we implemented an HPV vaccination social media promotion campaign at scale in Bauchi state, Nigeria. The combined phase 1 and 2 campaigns reached over 1.1 million unique Meta accounts, generated more than 12 million 3-second views, and generated more than 3.1 million full views. Self-reported exposure, as expected, increased as the 2 campaigns were implemented, from some 69% at baseline (likely due to existing online content related to the HPV vaccine), to nearly 81% at endline. This suggests that the campaigns not only reached large numbers of people, but that there was increasing awareness and recall of the HPV vaccination promotion messages.

Quasi-experimental evaluation findings, based on the heavy-up design,^22^ suggest that the campaign was effective in increasing HPV vaccination rates based on a quasi-experimental evaluation. Specifically, in a longitudinal sample, there was an increase in vaccination rates associated with higher levels of engagement with the campaigns. Engagement was measured through social media metrics, and there were over 825,000 individual engagements (eg, likes, shares, comments) with posts across the phase 1 and 2 campaigns. However, we did not see a relationship between self-reported recall of campaign posts and higher vaccination rates.

Overall, in our sample, vaccination rates increased from 69% at baseline to 83% and midline and 86% at endline. We also found that the pro-vaccine attitudes factor, derived from factor analysis of a series of items that asked about and reduced vaccine hesitancy also was associated with higher vaccination rates. The relationship between positive vaccinated-related attitudes and subsequent vaccination has been examined in recent studies, with mixed results.^29^ Researchers have found an attitude-uptake gap in regard to HPV vaccination.^30^ Some studies, like ours, have found that substantial increases in pro-vaccine attitudes may be associated with increases in HPV vaccine uptake.^31^ But given variable results in such studies, there is no clear evidence base on the role of pro-vaccine attitudes, and related beliefs and intentions, in promoting uptake. This area, and how innovative methods such as social media campaigns to promote pro-vaccine attitudes and intentions, calls for more rigorous interventions and research.

Furthermore, we found that the campaign produced additional HPV vaccinations at low cost: Adjusted for campaign exposure cost of $0.81 per additional vax in campaign phase 1, $2.10 per vax in phase 2, and $2.40 for the combined campaigns. There is limited evidence on the effectiveness of social media campaigns to promote vaccination generally, and HPV vaccination specifically.^32^ However, the present study adds to previous research in LMICs that suggests social media is a promising strategy to increase vaccine uptake. More rigorous interventions, combined with cost effectiveness analysis, should be conducted to build the evidence base in this area.

Overall, evidence from this study suggests that social media may be an effective strategy to promote HPV vaccination, and that it is cost effective compared to alternatives. More research is needed on the mechanisms and determinants of HPV vaccine uptake, including the role of psychosocial variables such as vaccine hesitancy, pro-vaccine attitudes, and social norms.^33,34^ Additionally, rigorous study designs, including randomized controlled trials, and dose-response experiments of social media campaigns, are needed to help interpret campaign impact on HPV vaccine uptake and behavioral determinants.^35^ Such studies are also needed to build the evidence and allow for valid comparisons between studies on social media vaccination cost effectiveness analysis.

Overall, this research suggests that future interventions should utilize social media to promote vaccination, and that the types of campaign strategies used in this study may be worth using or adapting. These include use of storytelling narratives, humor, and messages delivered by trusted and influential voices (social influencers). While much more evidence is needed on these techniques, other studies have found them to be promising ways to frame health messages, and promote behavior change.^36^

Despite the evidence provided by this study, it has some important limitations. First, we experienced high attrition of approximately 75%. While attrition is often high in social media-based studies, this level of attrition means that our findings should be judged with caution. The evidence from this study, while promising, requires corroboration in trials with higher retention. Second, there were other social influences and media messages both pro-vaccination, and misinformation, related HPV during the time of our study. Efforts were ongoing in Bauchi, and across Nigeria, to promote HPV vaccine uptake, and also unmeasured negative influences. These may present important confounds to our study, and future research should examine their potential impact. Third, this was not a randomized study, so while the heavy-up design provides a mechanism for causal inference, it has limitations. For example, attention to and engagement with the campaign may be attributable to other factors that would influence the likelihood of HPV vaccine uptake. Future studies should use randomized designs. Finally, the nature of Meta’s social media metrics prevents individual level attribution of campaign exposure to outcomes. We have data on the exposure in the population, and can relate those metrics to our survey data, but not to individual study participants.

## CONCLUSION

Social media campaigns show promise for HPV vaccination promotion. There is a small, but growing body of evidence that social media can increase vaccination rates, and that higher levels of exposure to and engagement with such campaigns is associated with vaccine uptake. These campaigns show evidence of producing high vaccine uptake at low cost. Future research should use rigorous designs and examine specific factors (eg, the role of pro-vaccine attitudes and social norms) that may predict uptake.

## Data Availability

The minimal data set, and accompanying code, will be made available upon manuscript acceptance.

## ACKNOWLEDGEMENTS

This research was supported by Gates Foundation grant INV-070804.

## AUTHOR CONTRIBUTIONS

**Conceptualization:** W. Douglas Evans.

**Data curation:** W. Douglas Evans, Alec Ulasevich, Sean O’Sullivan.

**Formal analysis:** W. Douglas Evans, Alec Ulasevich.

**Funding acquisition:** Drew Bernard, Sarah Francis.

**Investigation:** W. Douglas Evans.

**Methodology:** W. Douglas Evans, Sarah Francis, Drew Bernard.

**Project administration:** W. Douglas Evans, Sarah Francis, Julie Szabo, Ibrahim Sani Mohammed Gimba.

**Resources:** W. Douglas Evans

**Software:** Alec Ulasevich.

**Supervision:** W. Douglas Evans.

**Validation:** W. Douglas Evans, Alec Ulasevich.

**Visualization:** W. Douglas Evans, Alec Ulasevich.

**Writing – original draft:** W. Douglas Evans, Alec Ulasevich.

**Writing – review & editing:** W. Douglas Evans, Alec Ulasevich, Sarah Francis.

## Notes

### Competing Interest Statement

The authors have declared no competing interest.

### Author Declarations

The National Health Research Ethics Committee (NHREC) of Nigeria and the Bauchi State Ministry of Health reviewed and approved the study protocol.

